# Machine learning based prediction of antibiotic sensitivity in patients with critical illness

**DOI:** 10.1101/19007153

**Authors:** Debabrata Ghosh, Shivam Sharma, Eeshan Hasan, Shabina Ashraf, Vaibhav Singh, Dinesh Tewari, Seema Singh, Mudit Kapoor, Debarka Sengupta

## Abstract

Rising antibiotic resistance inflicts a heavy burden on healthcare, both clinically and economically. Owing to the time required to obtain culture and sensitivity test results, quite often the clinicians rely on their experience and static clinical guidelines to prescribe antibiotics. Such empirical treatment often fails to account for patient-specific attributes and changes in the antibiotic resistance patterns with time and location. The aim of this study was to analyze the patient and hospital specific features regarding their prognostic relevance to treat bacterial infections of patients in the intensive care units (ICUs). We performed a single-center retrospective cohort analysis across 25526 positive cultures recorded in MIMIC-III critical care database. We retrieved a number of clinically relevant relationships from association analysis between patient factors and bacterial strains. For instance, higher elapsed time from patient admission to sample collection for culture showed strong association with blood stream infection caused by *Enterococcus faecium, Pseudomonas aeruginosa*, and *Staphylococcus*, indicating that these infections are possibly hospital acquired. To predict antibiotic sensitivity at the level of individual patients we developed an ensemble of machine learning algorithms. The model provided superior prediction accuracy (about 87%) and area under the ROC curve (around 0.91 on an average) for the four most common sample types as compared to a number of off-the-shelf techniques. We demonstrate the predictive power of commonly recorded patient attributes in personalised prediction of antibiotic efficacy.

## Introduction

The aggravating loss of antibiotic activity as a result of the dispersal of resistant genes among micro-organisms has become a global health challenge and a threat to humankind. Antibiotic resistance inflicts a heavy burden on healthcare both clinically and economically, with 23,000 and 25,000 estimated annual deaths respectively in the United States and in Europe as well as increasing length of stay and morbidity. Reports suggest a projected annual death toll touching ten million worldwide by 2050^1–3^. In the United States alone, the annual cost associated with antimicrobial resistance has been estimated to be $55 billion^4^. Intensive Care Units (ICUs) in hospitals are often considered to be the epicenter of development, acceleration, and proliferation of drug-resistant microorganisms^5, 6^. Critically ill patients in ICU are particularly vulnerable to infections due to their exposure to multiple invasive procedures. These include mechanical ventilation, tracheal intubation, vascular access etc. which, in turn, leads to compromised defense mechanism of anatomical barriers, impairment of protective mechanisms such as cough reflex by sedative drugs, and the frequent impairment of the immune response induced by trauma, surgery, and sepsis^7^. According to a dated multi-center study (conducted in 1992) in Europe, around 20% to 30% of ICU admissions had reported an incidence of nosocomial infection^8^. A more recent study (conducted in 2007) involving 1265 ICUs from 75 countries reported the presence of hospital-acquired infections in about 50% of ICU patients^9^.

In today’s world of rapidly growing antimicrobial resistance (AMR), optimal antibiotic prescription is crucial in critical care settings^10^. Frequently used broad-spectrum antibiotics are among the primary drivers of rising AMR. Reports suggest that 30% to 60% of the antibiotics prescribed in ICUs are unnecessary, inappropriate, or suboptimal^11–13^. Epidemiological studies have demonstrated a direct relationship between antibiotic consumption and the emergence/propagation of several resistant strains in ICUs^11–14^. Nosocomial infections, caused by multi-drug resistant (MDR) organisms, are more prevalent in ICUs as compared to other departments^8^. Infection caused by MDR organisms often result in much worse clinical outcomes compared to their susceptible counterparts^15^. Such outcomes have also been linked with delay in the administration of right antibiotics^16^.

In the presence of clinical symptoms, detection of the pathogen via culture remains the gold standard for diagnosing the majority of the bacterial infections including urinary tract infection (UTI) and bloodstream infection (BSI)^16–18^. Culture and Sensitivity (C & S) report provides definitive evidence of a particular infection in a subjected specimen^19, 20^. C & S reports also cite antimicrobial susceptibility of the individually tested drugs through Minimum Inhibitory Concentration (MIC) values. Generation of a C & S report typically takes around 24 to 72 hours. In its absence, physicians rely on their perception about the clinical presentation of the specific cases and other available clinical guidelines^21^. Such approaches could potentially disregard patient-specific attributes and the temporal changes in the antimicrobial resistance patterns^22^. Appropriate choice of empiric antibiotic must balance out the objective to minimize the prescription of broad-spectrum antimicrobial agents and give a broader spectrum of coverage across various bacterial strains^23^. It has been shown that inappropriate empiric therapy is associated with poorer outcomes with longer length of stay, increased health-care costs, higher morbidity, and mortality^24^.

Numerous studies have suggested appraisal of pathogen etiology in a localized setting as the ideal basis of empiric therapy^25–27^. We performed a retrospective study to correlate antibiotic resistance to a broad range of patient-specific factors such as gender, comorbidities, site of infection, the events of past hospitalization, and previous antibiotic usage. These factors show significant non-monotonic associations with the efficacy of the antibiotics. We used patient information and culture data of 11496 patients from the Medical Information Mart for Intensive Care III (MIMIC-III, data collected between 2001 and 2012) critical care database^28^. We first analyzed the bacterial prevalence and susceptibility patterns across bacteria-antibiotic pairs. Further, we presented a statistical approach unraveling the complex landscape of association between various patient-related features and the bacterial species. This analysis provided us with a comprehensive set of clinically relevant relationships. Finally, we explored the potency of various concerned patient factors to predict the susceptibility of bacteria to specific antibiotics. To this end, we developed an ensemble of machine learning models for personalized antibiotic susceptibility prediction that yielded a high overall accuracy, compared to some of the existing best practice methods.

## Results

### Overview of the data

MIMIC-III is a publicly available database consisting of de-identified Intensive Care Unit (ICU) patient data from Beth Israel Deaconess Medical Center (BIDMC) in Boston, Massachusetts from 2001 to 2012^28^. This comprehensive healthcare data includes detailed information of patients’ demographics, admission details, laboratory tests, vital signs, medications, microbiology procedures, and mortality to name a few. From the total C & S reports of 11496 patients, we only considered the positive cultures for the current study. Subsequently, we found 27625 unique culture isolates. We illustrated the bacterial distribution and susceptibility patterns for most frequently occurring bacteria including *Staphylococcus aureus, Escherichia coli, Enterococcus*, and *Pseudomonas aeruginosa* in Fig 1a and Fig 1b respectively. We mainly considered the top four types of sample for this study. We found that the most frequently collected four samples were sputum, urine, blood, and pus swab. From the diagnoses table, we captured 18 different comorbidities namely ‘endocrine, nutritional and metabolic diseases, and immunity disorders’, ‘diseases of the blood and blood-forming organs’, ‘mental disorders’, ‘diseases of the nervous system and sense organs’, ‘diseases of the circulatory system’, ‘diseases of the genitourinary system’, ‘symptoms, signs, and ill-defined conditions’, ‘external causes of injury and supplemental classification’, ‘infectious and parasitic diseases’, ‘diseases of the musculoskeletal system and connective tissue’, ‘injury and poisoning’, ‘neoplasms’, ‘diseases of the respiratory system’, ‘diseases of the skin and subcutaneous tissue’, ‘diseases of the digestive system’, ‘congenital anomalies’, ‘certain conditions originating in the perinatal period’, and ‘complications of pregnancy, childbirth, and the puerperium’ while excluding ‘Factors Influencing Health Status And Contact With Health Services’. We showed the distribution of four specimens mentioned above along with other samples combined in Fig 2a. We regarded each admission a separate instance and plotted the distribution of the 18 aforementioned comorbidities along with cases with no comorbidity associated in the patient demography in Fig 2b. We gave the detailed description of variables retained after extraction and feature engineering in Table 1.

**Table 1.** Detailed description of variables extracted and created for retrospective analysis. Patient demographic, clinical conditions, admission details and culture details across all positive cultures were collected which comprises of several categorical, continuous and integer-valued variables.

| Variables |  | Variable Type and Description |  |
| --- | --- | --- | --- |
| **Demographic** | Gender | Categorical/Nominal<br>(2 categories) | Male: 12650 samples<br>Female: 10414 samples |
|  | Age | Continuous | Computed at the time of admission |
| **Admission Details** | Number of Previous Admissions | Integer-valued | Number of times patient was previously admitted before current admission |
|  | Sample Collection Interval | Integer-valued | Interval in days between admission and sample collection |
| **Patient Condition** | Existing Comorbidities | Categorical/Nominal | No comorbidity associated or one of 18 different comorbidities among ‘endocrine, nutritional and metabolic diseases, and immunity disorders’, ‘diseases of the blood and blood-forming organs’, ‘mental disorders’, ‘diseases of the nervous system and sense organs’, ‘diseases of the circulatory system’, ‘diseases of the genitourinary system’, ‘symptoms, signs, and ill-defined conditions’, ‘external causes of injury and supplemental classification’, ‘infectious and parasitic diseases’, ‘diseases of the musculo-skeletal system and connective tissue’, ‘injury and poisoning’, ‘neoplasms’, ‘diseases of the respiratory system’, ‘diseases of the skin and subcutaneous tissue’, ‘diseases of the digestive system’, ‘congenital anomalies’, ‘certain conditions originating in the perinatal period’, and ‘complications of pregnancy, childbirth, and the puerperium’ |
|  | Device Inserted | Categorical/Nominal | No device inserted or one of 9 different devices (Hepatic Hunt Tips, CVL, Chest Tube, Abscess CHG, Catheter, Thoracostomy Tube, ABD Port, PEG, Arterial Line) inserted |
| **Culture Details** | Type of Sample sent | Categorical/Nominal | 27 types of specimen sent for culture |
|  | Organism present | Categorical/Nominal | 115 different organisms |
|  | Antimicrobial tested | Categorical/Nominal | 30 different antimicrobial tested |
|  | Bacterial sensitivity | Categorical/Nominal | Sensitive (S): 1313845 tests<br>Resistant (R): 546858 tests<br>Intermediate (I): 73533 tests |

**Figure 1.**
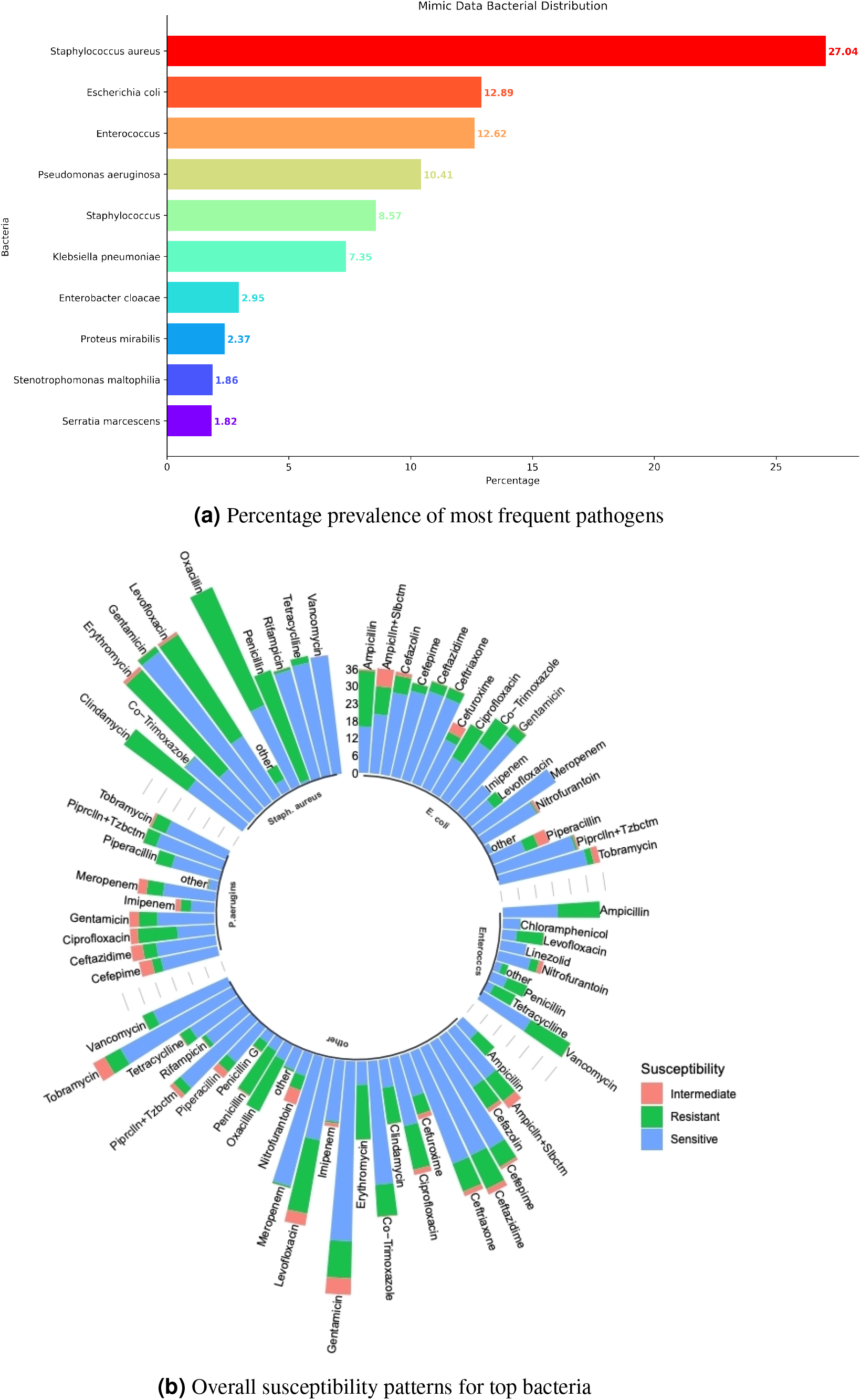
Bacterial distribution and susceptibility patterns for topmost bacteria. The percentages of prevalence for top 10 bacteria have been shown in Fig 1a and Fig 1b corresponds to a sunburst plot displaying overall susceptibility patterns of topmost few bacteria.

**Figure 2.**
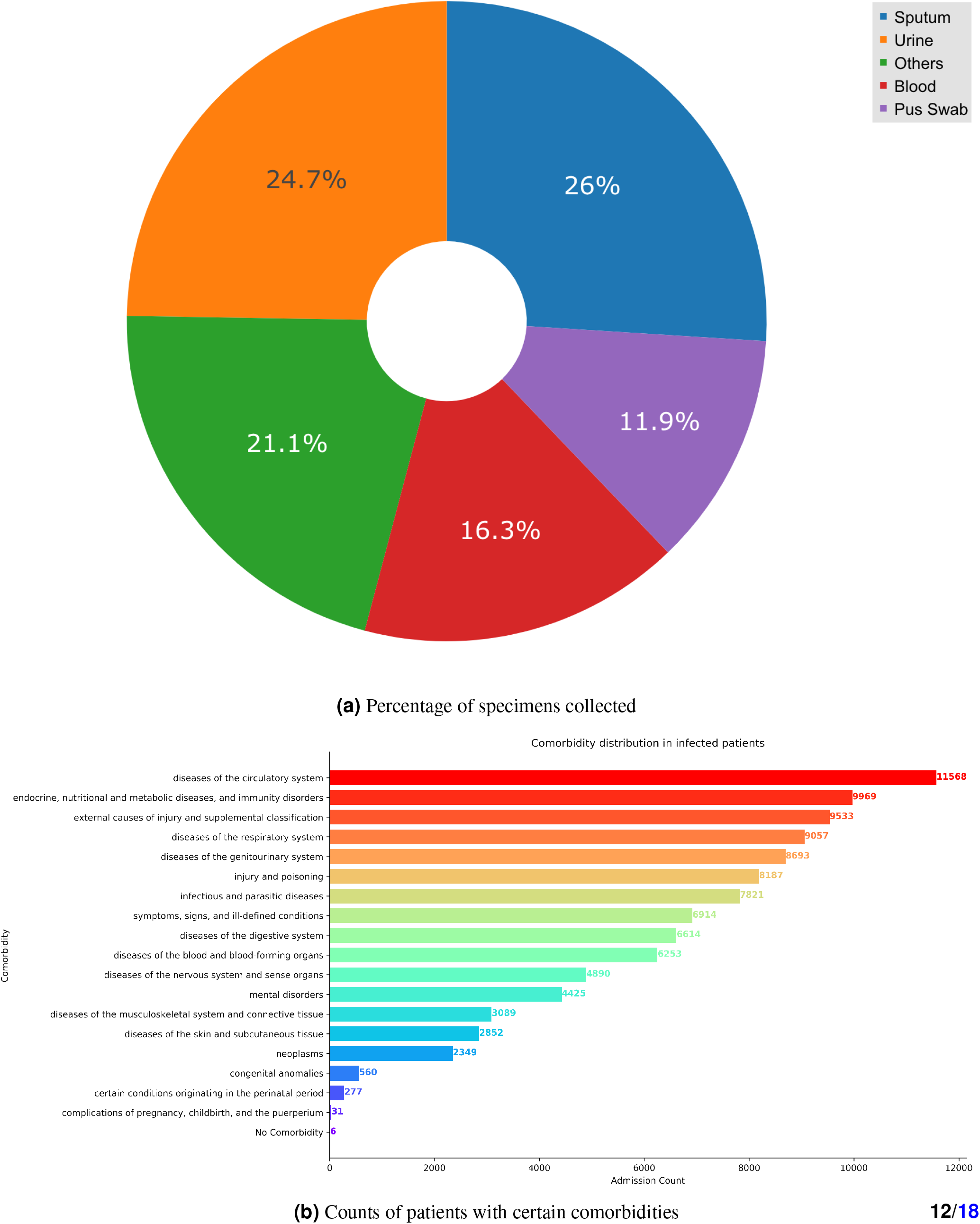
Distribution of specimens collected and comorbidities associated in patient population. Fig 2a shows the percentages of particular frequently collected samples or specimens and Fig 2b shows count of patient admissions with specific comorbidities.

### Antibiotic resistance patterns

We focused primarily on BSI cases and illustrated the heatmap of resistance patterns for the same in Fig 3. According to the data of blood samples, except for extensively used drugs such as penicillin (15.64% sensitive cases), oxacillin (28.4% sensitive cases), ampicillin (46.28% sensitive cases), erythromycin (27.24% sensitive cases), clindamycin (44.87% sensitive cases), levofloxacin (42.51% sensitive cases) most of the drugs used were quite effective in terms of bacterial sensitivity. Carbapenems such as imipenem and meropenem worked effectively with 96.24% and 96.36% sensitive cases against gram -ve bacteria. Notably, in 100% of the cases (total 1739) different strains of *Staphylococcus* species turned out to be sensitive to vancomycin. We attached the heatmaps of resistance patterns for all infection data in Figure S3 (Supplementary Methods) and other frequently occurring infections in Figure S4 (Supplementary Methods). There we clearly observed differences in bacterial prevalence as well as antibiotic-bacteria pairwise resistance rates for individual sample and entire data.

**Figure 3.**
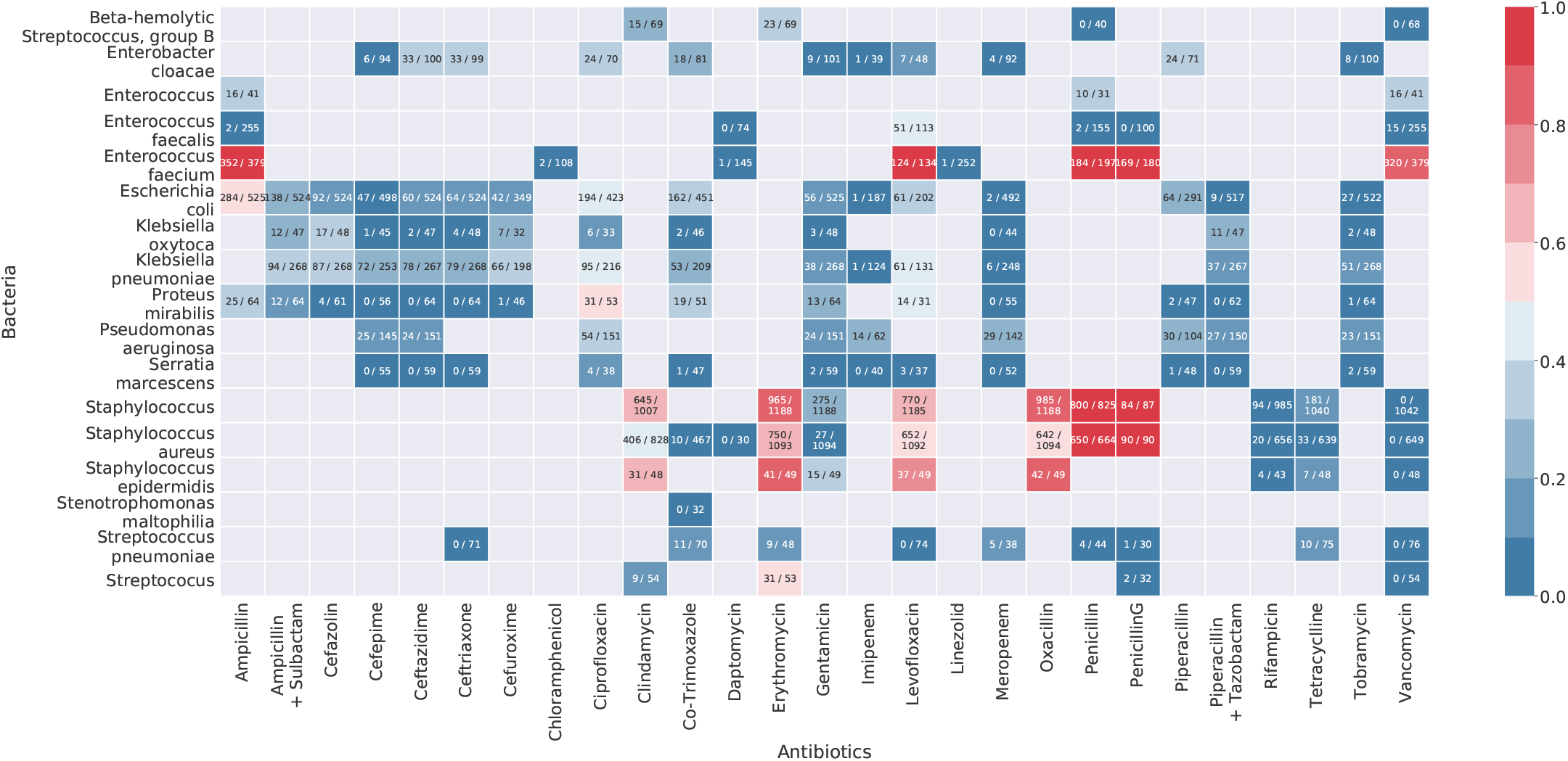
Antibiotic vs bacteria pairwise resistance rate in MIMIC-III data for blood sample. Each entry in the matrix denotes the ratio of number of resistance cases over total number of cases when the corresponding antibiotic in the column is tested against the bacteria in the row. The blank vacant entries refer to the cases when the antibiotic is never tested against the bacteria as per the data.

### Factors influencing bacterial infections

In the current study, we explored the association of different patient-related factors with the bacterial strains by using multinomial logistic regression for blood sample data. We presented the coefficients of regression in Fig 4. We reproduced the results of similar analysis for sputum, urine, and pus swab sample data in Figure S5 (Supplementary Methods).

**Figure 4.**
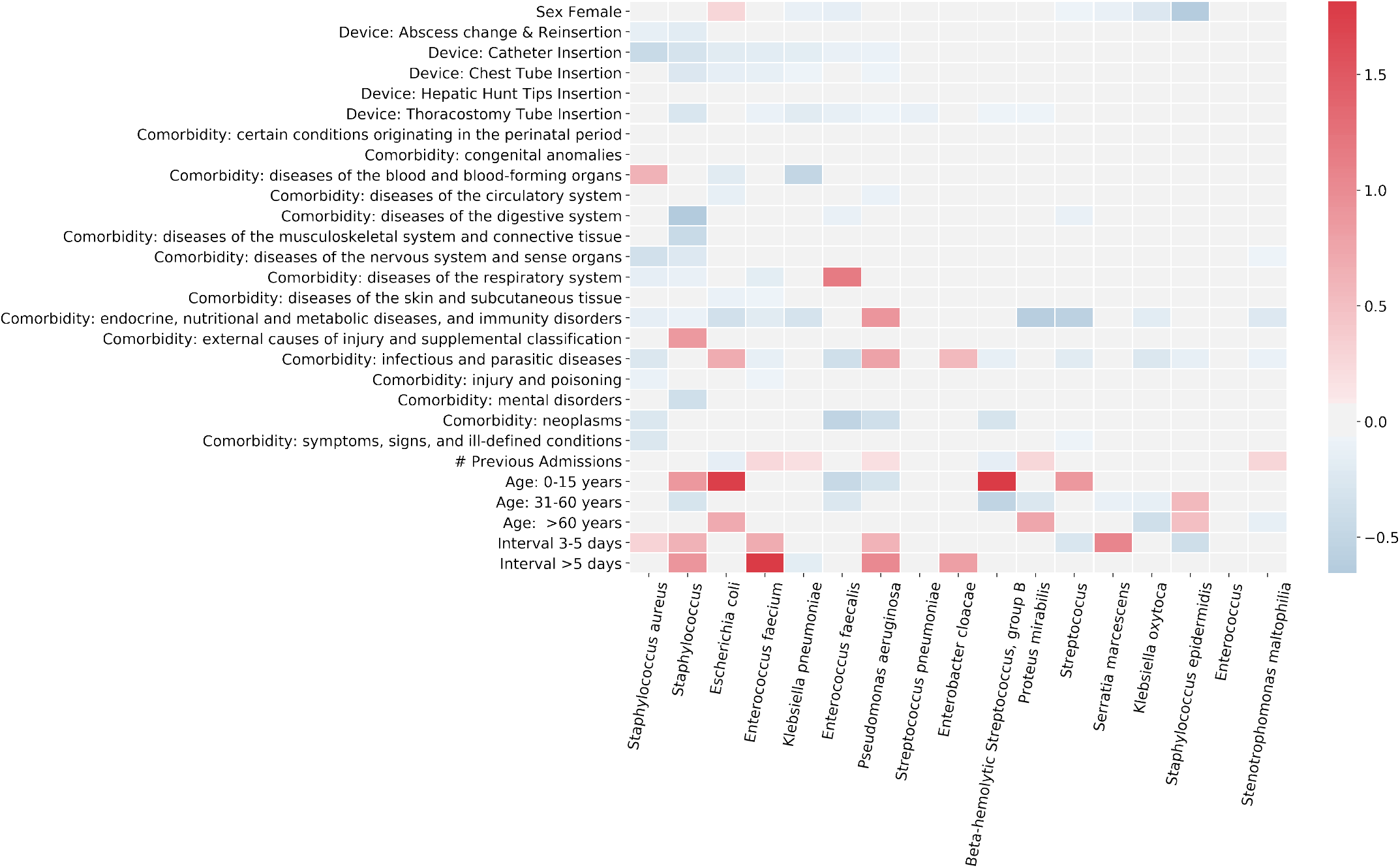
Coefficient plot of multinomial logistic regression on bacteria over several factors in blood sample data. The darker the red color gradient, the stronger the positive association between the corresponding bacteria and explanatory variables. Similarly with darker blue shade, stronger negative correlation is illustrated. The greyed out boxes implicates statistically insignificant associations.

We observed that *Enterococcus faecium* and interval of more than 5 days had the highest positive association with mean coefficient value ∼1.84. Next, we noticed that *Beta-hemolytic Streptococcus, group B* and *Escherichia coli* are more likely to cause infections in patients of age 0 − 15 years as compared to patients belonging to other age groups with mean coefficients 1.75 and 1.74 respectively. The bacteria being *Enterococcus faecalis*, ‘diseases of the respiratory system’ had a positive coefficient ∼1.17 on an average. Furthermore, 3 − 5 days after a patient’s admission into hospital, *Serratia marcescens* was found to have a higher infection rate, represented by the strong positive mean coefficient value of 1.04. We also noted that incidents of *Pseudomonas aeruginosa* were positively correlated with interval greater than 5 days and ‘endocrine, nutritional and metabolic diseases, and immunity disorders’ with mean coefficients 1.02 and 0.93 respectively. *Staphylococcus* also had positive association (mean coefficient value ∼ 0.92) with interval more than 5 days. On the other hand, *Staphylococcus* had strong negative correlation (mean coefficient value −0.66) with ‘diseases of the digestive system’. *Staphylococcus epidermidis* infections were much less frequent in females compared to males supported by mean coefficient value 0.63. We put the complete list of regression coefficients down in Table S1 (Supplementary Methods).

### Personalized prediction of antibiotic resistance

We tested the performance of our proposed approach with the demographic, clinical, and microbiology data extracted from MIMIC-III database after data cleaning, variable selection and baseline model choice. We randomly split the data of each type of specimen namely urine, sputum, blood, and pus swab into 70% for training and 30% for testing repeatedly for 100 times and calculated means and standard deviations of different performance measures to evaluate our model (Fig 5). We compared the performances of different models using metrics such as accuracy (Fig 5a), precision, recall, F1 score (Fig 5b), Cohen’s kappa score (Fig 5c), area under curve (AUC) of receiver operating characteristic (ROC) (Fig 5d) to name a few. We gave the complete comparison of different baseline models and our ensemble model in Table S2 (Supplementary Methods). We observed that our ensemble model outperformed the other models experimented in almost all aspects. We achieved an average accuracy of 85.8%, 87.03%, 86.28%, and 89.17% over 100 iterations for urine, sputum, blood, and pus swab samples respectively. The next best model for those samples attained 83.59%, 86.11%, 84.93%, and 88.82% accuracy respectively. We also yielded mean kappa scores of 0.58 (urine), 0.71 (sputum), 0.69 (blood), and 0.77 (pus swab). Mean AUC values for ROC corresponding to urine, sputum, blood, and pus swab samples came out to be 0.88, 0.92, 0.92, and 0.94 respectively.

**Figure 5.**
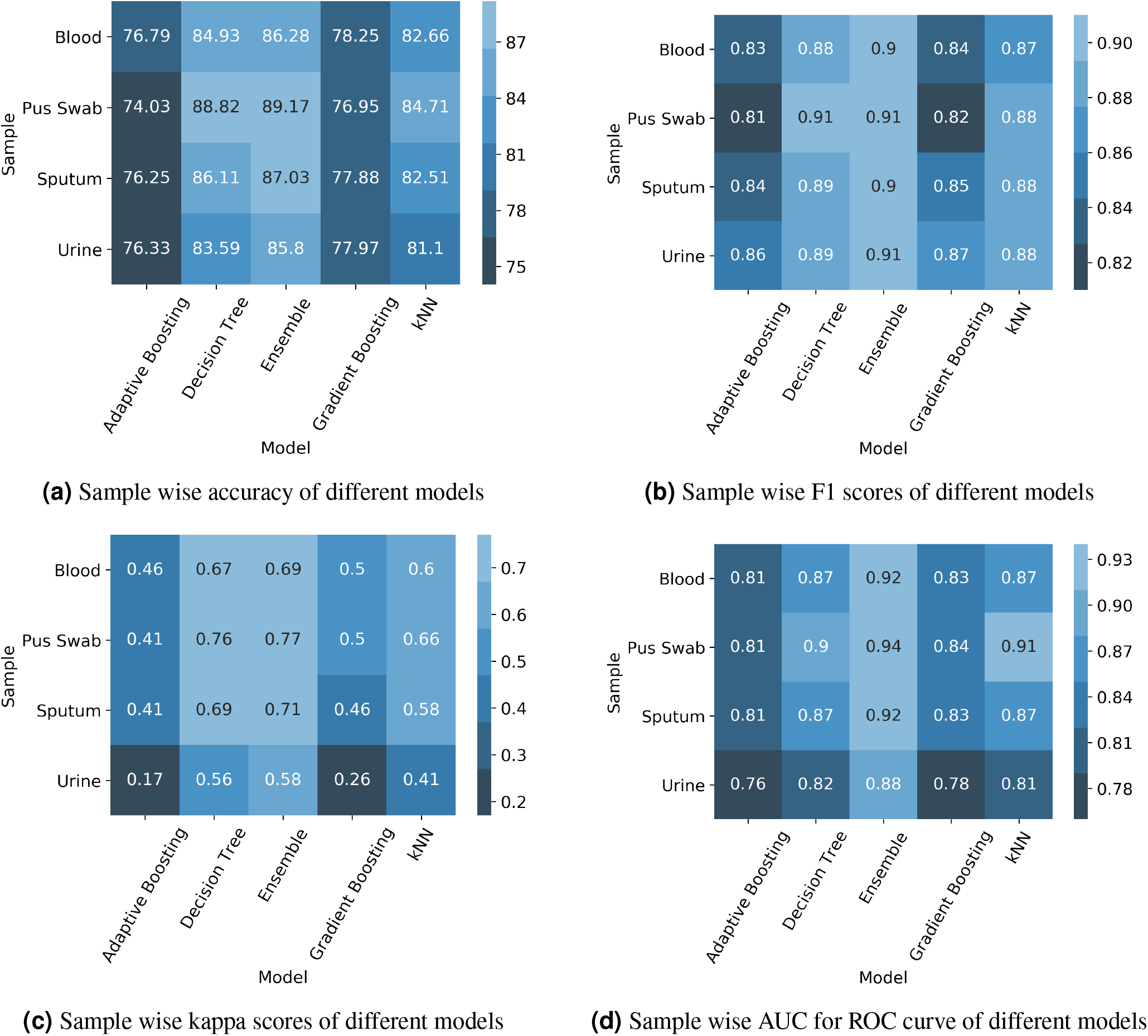
Performance measures for different classifiers over 100 iterations of 70-30 random split. Accuracy, F1 score, kappa score, and area under curve (AUC) for receiver operating characteristics (ROC) were calculated for all five top performing models: ensemble, decision tree, kNN with Hamming distance, Adaptive Boosting (AdaBoost) and Gradient Boosting on randomly split 30% test data. Fig 5a, Fig 5b, Fig 5c, and Fig 5d show sample wise accuracy, F1 score, kappa score, and AUC respectively.

We also plotted the ROC curve after splitting the data randomly into 70% and 30% for training and testing as shown in Fig 6 for four most prevalent sample types namely urine, sputum, blood, and pus swab. Our model yielded AUC values of 0.88, 0.93, 0.93, and 0.95 for urine, sputum, blood, and pus swab samples respectively in those instances.

**Figure 6.**
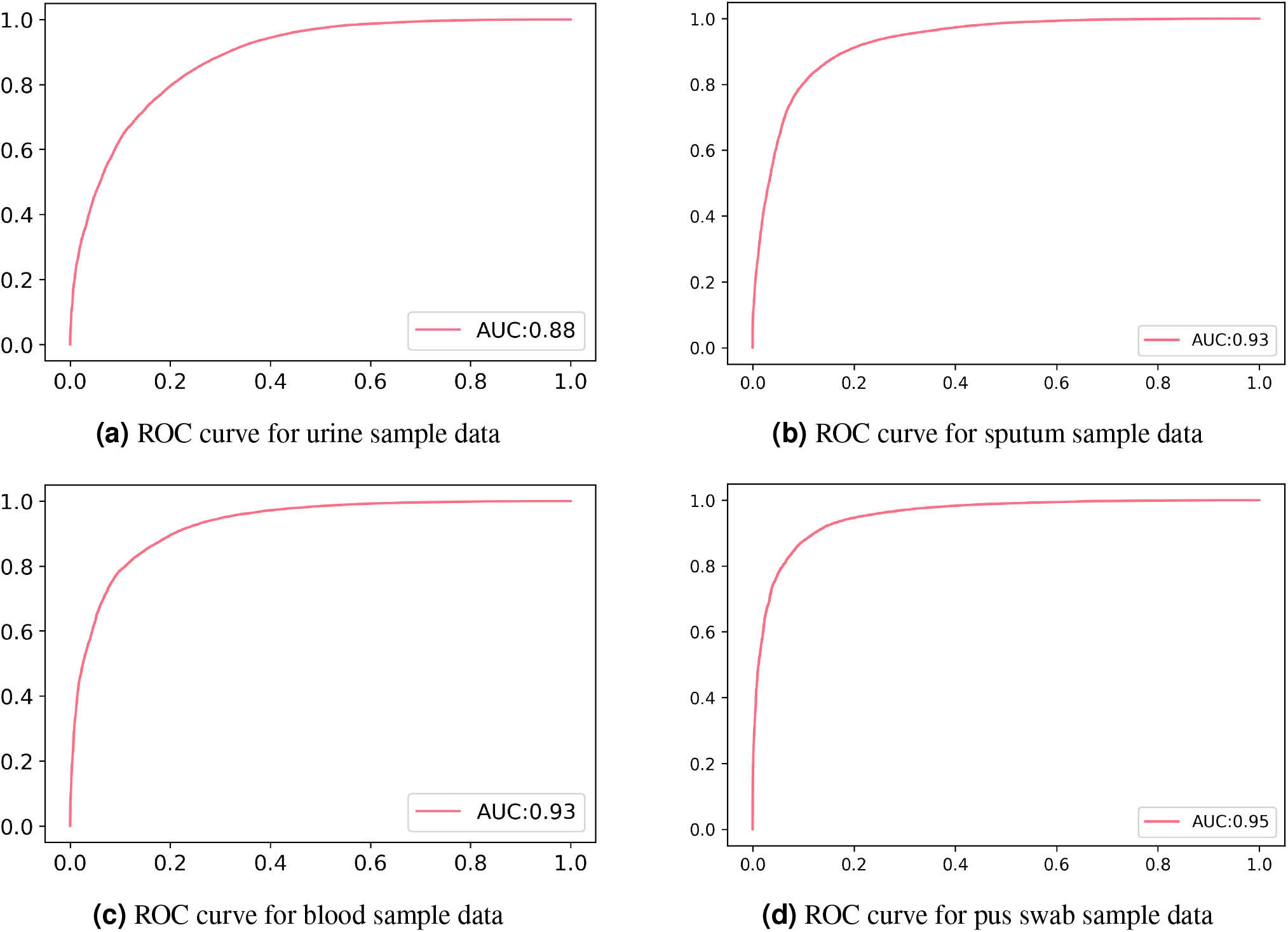
Sample specific ROC curves for ensemble model with 70-30 split. For our ensemble model with 5 base learners and gradient boosting meta learner, the receiver operating characteristic curve was plotted by training on randomly split 70% training dataset and testing on the remaining 30% test dataset. Fig 6a, Fig 6b, Fig 6c, and Fig 6d show sample wise ROC curves with AUC values for urine, sputum, blood, and pus swab respectively.

## Discussion

Advances in machine learning, coupled with training on large electronic health records and hospital integrated system datasets, have the ability to disrupt the areas of diagnosis and prognosis in emergency medicine^29,30^. In the space of bacterial infection, retrospective analyses of etiology and descriptive studies have been reported in regular intervals^31–33^. However, collective and methodical applications of predictive modeling is yet to be extensively implemented in the field of empiric treatment via antimicrobial sensitivity analysis barring a few interesting attempts^34,35^. Also the data used in the existing studies were not publicly available. Therefore, we could not validate their claim or compare our approach against the models applied in such studies. We looked at a wide range of variables comprising patient conditions, demographic information, previous culture history to predict susceptibility of specific bacterial species against antibiotics at a personalized level. The indiscriminate use of antibiotics has evidently resulted in increasing resistance of bacteria and subsequently rendered many of the standard treatment options ineffective. Rapid emergence of new strains of resistant bacteria made treatment of infections and monitoring of bacterial resistance substantially unpredictable. Inherent delay associated with producing C & S reports often prompts the doctors to treat the patient empirically without processing any culture. Also, in many developing and backward countries cultures are not ordered considering the economic burden of negative cultures. Our aim is to bring down inappropriate prescription and cognitive load of the physicians in such scenario. By means of heatmaps showing bacteria-antibiotic pairwise resistance pattern and relative bacterial prevalence given patient factors, data visualization can help physicians inferring bacterial prevalence and their corresponding resistance not only in care type or ward level (e.g. ICU) but also in patient level. The proposed data-driven antimicrobial susceptibility test (AST) can be used to generate highly accurate patient-specific predictive antibiograms. In absence of the culture report, these predictive antibiograms can aid clinicians in complex, data-backed decision making while recommending antibiotics.

Blood stream infection is one of the most critical and prevalent infectious diseases. Moreover, BSI management is often suggested as a performance indicator for antimicrobial stewardship programs in healthcare facilities. Therefore, we chose to perform most of our statistical analysis using blood sample data only. We developed a novel statistical approach to discover the association between patient factors and incidence of infections caused by specific bacterial species. We found several interesting clinically relevant relationships, which corroborate or enhance our present understanding of the causes of infections. These associations can be used by antibiotic stewardship teams to implement hospital-specific guidelines in a scientific manner. Although we observed that for a large number of patient factors, we could not infer any association. In such a scenario, the presence of additional data might allow us to make more inferences that are statistically significant. Moreover, identifying the presence of a bacterial strain in patient level might help stewardship team to track the bacterium with more granularity. This will in turn help them implementing measures to curb the spread of infection within the hospital against present alarming situations as reported by David and colleagues.^36^ We observed that infections caused by *Enterococcus faecium, Pseudomonas aeruginosa*, and *Staphylococcus* are more common in patients with increased interval between admission and sample collection dates, indicating that these infections are probably hospital acquired. Past studies also suggested that bloodstream infections due to *Pseudomonas aeruginosa* and *Enterococci* are usually hospital acquired^37^. According to our analysis, *Escherichia coli* and *group-B streptococcus* infections were prevalent in patients of 0−15 years of age. Earlier investigations mentioned infection due to *group-B streptococcus* and *Escherichia coli* as a major cause of neonatal sepsis^38^. This corroborates our findings.

With the off-the-shelf machine learning models we did not do particularly well except for decision tree in terms of performance and calibration. Therefore, we formulated a new approach which gave us a significant delta jump in accuracy from the existing classifiers yielding approximately 87% accuracy on an average across all test cases. Receiver operating characteristic (ROC) curve is a graphical representation of true positive rate (TPR) or sensitivity and false positive rate (FPR) or (1 - specificity) on y and x axes for varying cut-off points of test values. The area under the curve (AUC) is an effective measure of model validity which lies between 0 and 1 with a higher score indicating better performance. Therefore we concluded that the overall performance of the custom ensemble model on the test sets of different types of specimen (e.g. ROC-AUC in case of blood sample data: 0.92) was better than either of decision tree, kNN, gradient boosting, and AdaBoost (corresponding AUC: 0.87, 0.87, 0.83, and 0.81 respectively). Our model was able to calibrate to different prevalence of bacteria and their corresponding susceptibilities (sensitive or resistant) throughout the hospital for different infection types in the bootstrapped test sets.

Still, our current model has several limitations in terms of generalization, prospective study and variable selection. Our model is trained on the data from a single hospital which is not validated to be generalizable to multiple healthcare settings. The database did not contain any information about nosocomial infections or other factors contributing to the healthcare associated infections such as intubation or ventilation leading to infection. Documentation of whether the patients had religiously taken the prescribed antibiotics in the duration of their treatment was not available too. Even with a handful of commonly recorded patient information, our model was found to be accurate enough for being considered as a decision support system. As a continuation of our study we plan to prospectively examine the impact of our current model on future predictions and reduction of diagnostic error to subsequently improve the efficacy. We will also evaluate the effectiveness of the predictive power of our model in reducing inappropriate prescriptions and antibiotic consumption. We also intend to utilize bacteria-antibiotic pair susceptibility patterns in reference laboratories for a more comprehensive study in community acquired infections.

## Methods

### Extraction of relevant variables

For this study, we collected the data generated by microbiology laboratory from MICROBIOLOGYEVENTS table in MIMIC-III database along with the patients’ demographic and admission details from PATIENTS table and ADMISSIONS table respectively. We merged the table containing microbiology results for the patients detected with positive cultures and the table with patient details through a left outer join. Patient data obtained from the three tables mentioned can be categorized as follows: a. De-identified Patient identity - patient id, admission id; b. Patient demography - age and gender; c. Admission details - admission date, admission type; d. Patient condition - existing comorbidities as per the diagnosis, device information (e.g. Catheter insertion). For the cases with no reported comorbidity, we maintained a separate category and same for patients with no device associated before sample collection. Culture-related details extracted from the MICROBIOLOGYEVENTS table consisted of the name of the specimen or sample sent for culture, date and time of sending the sample for culture, the name of the bacteria grown on culture, antibiotic name, and susceptibility to the tested antibiotics. We derived few other variables from the available patient information including the number of previous admissions, the interval between admission and sample collection in days. For capturing the comorbidities, we used the DIAGNOSES_ICD table of the MIMIC-III database. We extracted the ICD-9 codes available for each admission as identified by patient id and admission id. We eventually ended up with 6984 unique ICD codes with quite sparse data. Numbers of comorbidities associated with each admission ranged from 0 to 39 while the average was 11. To tackle the sporadic feature of comorbidities associated, we only extracted the first three digits of the ICD codes for all except ‘Supplementary Classification Of External Causes Of Injury And Poisoning’ where the codes start with an ‘E’^39^. We then further merged them into 19 different categories as given in [ICD-9 data website](http://www.icd9data.com/2012/Volume1/default.htm).

### Resistance pattern across bacteria and association analysis of bacteria

For each bacteria-antibiotic pair, we computed the rate of resistance on the entire data. Since the bacterial prevalence can vary across infection sites and sample types, we also performed the same pairwise analysis for each type of sample. In overall and sample wise analyses, we only retained the bacteria-antibiotic pairs with at least 30 tests available on entire data and sample specific data respectively.

We used multinomial logistic regression to calculate the relationship between bacteria and the explanatory variables (patient data). Multinomial Regression uses a softmax estimator to estimate the probability of each class (see Eq (1)). We denote *x*^(*i*)^ to be the vector of predictors and *Y* ^(*i*)^ to be the bacteria for the *i*^*th*^ patient sample. The coefficient vectors *θ*_*k*_s capture the relationship between the set of input variables and the probability of observing an infection due to bacteria *k*. We obtained the coefficient vectors using *sklearn*.*linear*_*model*.*LogisticRegression* method, which employs *newton* − *cg* as the solver.

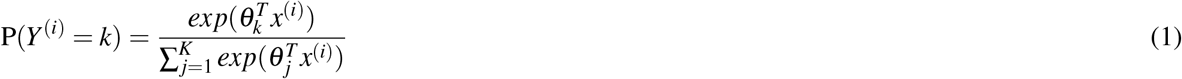

We computed the 95% confidence intervals of the coefficients by using *bootstrap*. If the confidence intervals included zero, we refrained from concluding any association. In the remaining cases, we reported either positive or negative associations. We did this analysis for each type of sample separately focusing primarily on blood sample.

### Prediction of antibiotic sensitivity at patient level

We used a stacked ensemble method to predict patient-specific antibiotic susceptibilities. We tried to capture as many different kinds of baseline models as possible to build the ensemble so that we can generalize and enhance model performance. However addition of each new model is associated with increased time and structural complexity. So we tried to minimize choosing models with correlated prediction errors. We started with tree-based models such as decision tree and random forest, boosting algorithms like adaptive and gradient boosting as well as other diverse classifiers namely k-nearest neighbors (kNN), naive bayes, and logistic regression. We greedily picked the models whose prediction errors are relatively uncorrelated on an average with other models. We illustrated the correlation matrix of prediction errors for all base learners in Fig 7 corresponding to blood sample data. As evident from the correlations shown in Fig 7, we greedily picked up decision tree, gradient and adaptive boosting, naive Bayes, and kNN as our baseline models for this case. We have added the correlation matrices of prediction errors corresponding to other specimens in Figure S1 (Supplementary Methods). Finally we used a gradient boosting classifier as a meta learner to combine the predictions of the selected base learners. The complete structure of the ensemble model for blood sample data is given in Fig 8.

**Figure 7.**
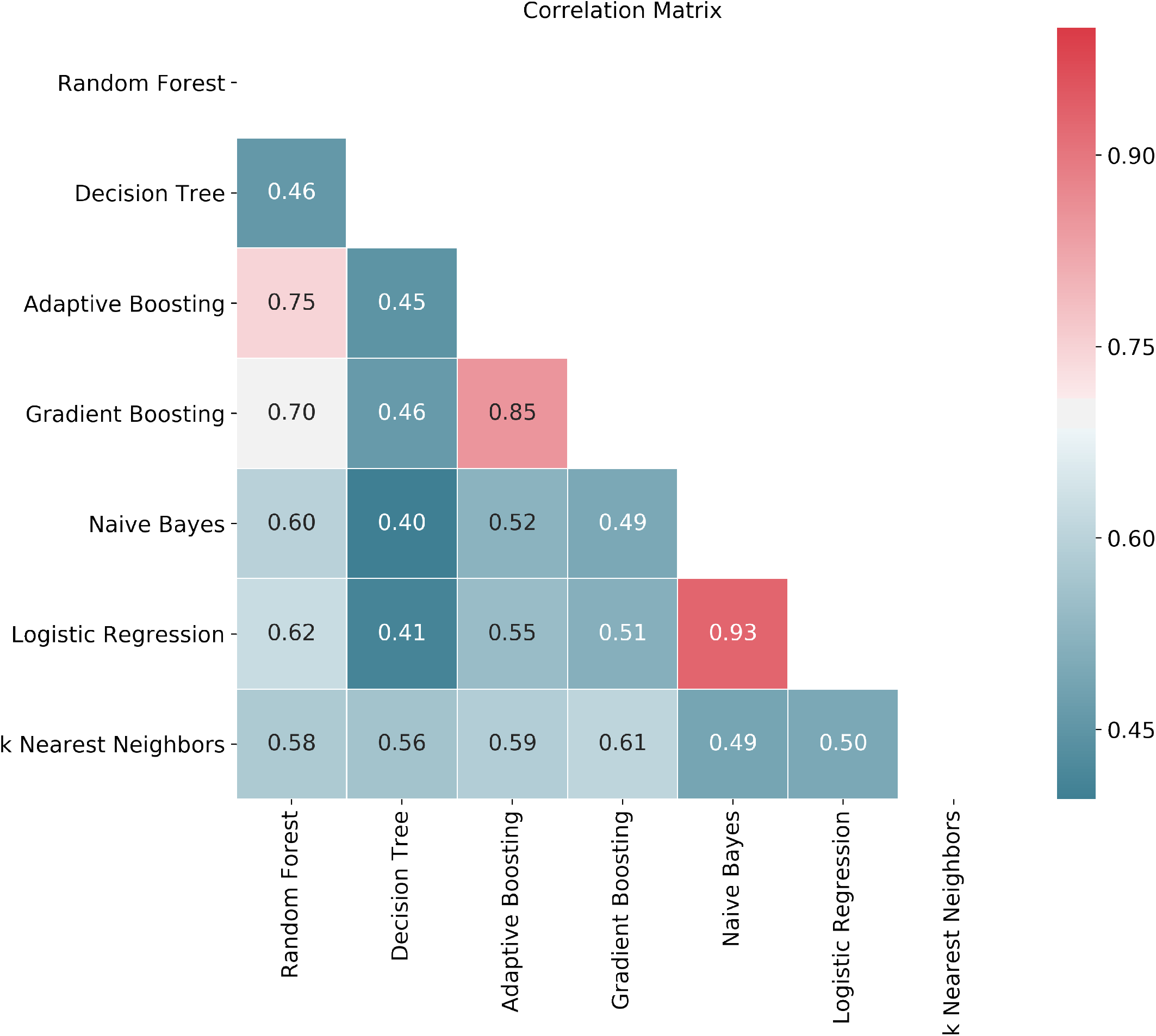
Correlation matrix of prediction errors for different baseline models corresponding to blood sample data. Prediction errors on randomly split 30% test data have been calculated for decision tree, adaptive boosting, gradient boosting, naive Bayes, logistic regression and kNN model for certain 70-30 data split to produce the correlation matrix.

**Figure 8.**
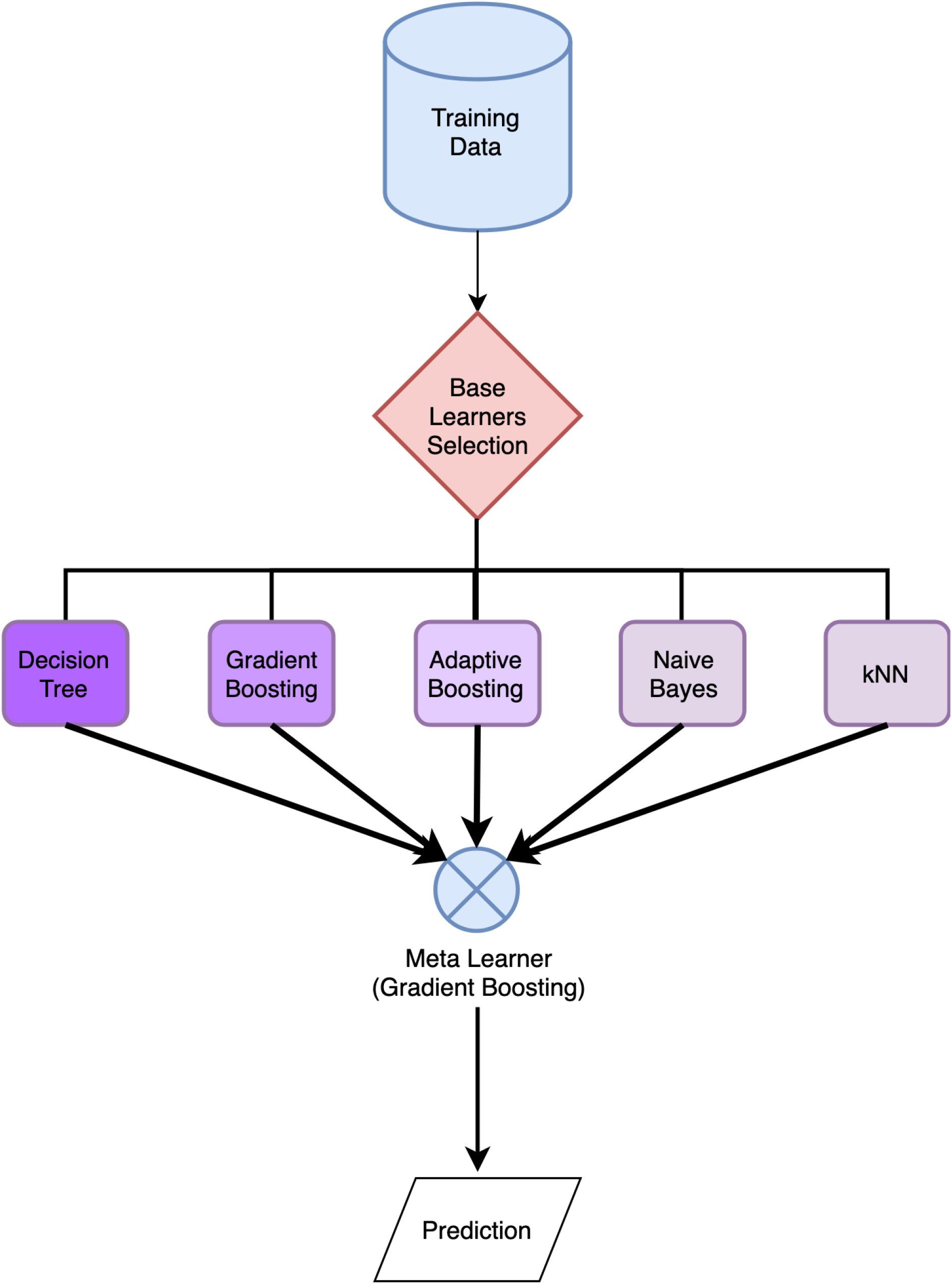
Complete structure of ensemble model. The baseline models were chosen focusing on minimizing prediction error correlation. Gradient Boosting classifier was used as a meta learner for combining the predictions of base learners.

Only data available during a patient admission until the time of culture along with previous admission information were used as prediction variables. To do away with missing or incomplete data, we did a complete case analysis. A set of 8 variables (age, gender, number of previous admissions, sample collection interval, list of comorbidities, devices inserted, specimen, and antibiotic) was selected through subject matter expert (clinician) knowledge and extensive literature review. We preferred expert opinion and literature review based selection over automated variable selection techniques to deal with accountability and user acceptance of features used in prediction. After preprocessing of raw data, we devised our binary classification algorithm to train our classifier for predicting susceptibility of the bacteria (resistant or sensitive) to the administered antibiotic. We classified intermediate levels of resistance as resistant to exclude possible inappropriate use of antibiotics with less efficacy. We executed this in a bacteria agnostic manner to use our prediction before sample is collected or sent for culture. Vis-à-vis, we created the data to be used for training and testing by label encoding and randomly splitting the data into 70% for training and 30% for held-out testing and evaluation. We repeated this process 100 times to obtain mean and standard deviation of different evaluation metrics for final model selection. We have attached the diagram of complete step wise procedure in Figure S2 (Supplementary Methods). We repeated the whole process for each type of specimen or sample and reported results for top four samples.

## Data Availability

All research has been carried out using publicly available MIMIC critical care database.

https://mimic.physionet.org/

## Acknowledgements

The study is supported by Circle of Life Healthcare Pvt. Ltd., New Delhi, India.

## Author contributions statement

D.S. conceived the project. D.G., S.S. (Shivam Sharma), E.H. and D.S. designed the experiments. D.G. implemented the ML algorithm and statistical analyses with help from S.S. and E.H. alike. The remaining authors contributed equally in conducting the experiments and analyses. All the authors discussed the results, co-wrote and reviewed the manuscript.

